# Optimal Graph Representations and Neural Networks for Seizure Detection Using Intracranial EEG Data

**DOI:** 10.1101/2024.12.28.24316703

**Authors:** Alan A. Díaz-Montiel, Richard Zhang, Milad Lankarany

## Abstract

In recent years, several machine-learning (ML) solutions have been proposed to solve the problems of seizure detection, seizure characterization, seizure prediction, and seizure onset zone (SOZ) localization, achieving excellent performance with accuracy levels above 95%. However, none of these solutions has been fully deployed in clinical settings. The primary reason has been a lack of trust from clinicians towards the so-called black-box decision-making operability of ML. More recently, research efforts have focused on explainability frameworks of ML models that are clinician-friendly. In this paper, we conducted an analysis of graph neural networks (GNN), a paradigm of artificial neural networks optimized to operate on graph-structured data, as a framework to detect seizures from intracranial electroencephalographic (iEEG) data. We employed two multi-center international datasets, comprising 23 and 16 patients and 5 and 7 hours of iEEG recordings. We evaluated four GNN models, with the highest performance achieving a seizure detection accuracy of 97%, demonstrating its potential for clinical application.

## I. Introduction

Epilepsy is a neurological disorder affecting over 50 million individuals worldwide [1], often necessitating thorough assessment in specialized units like the Epilepsy Monitoring Unit (EMU). Patients in these units undergo continuous video and electroencephalographic (EEG) monitoring, generating vast amounts of data—ranging from 1 to 10 terabytes per individual. Approximately 30-40% of these patients require intracranial EEG (iEEG) monitoring [2], [3], involving the implantation of electrodes within the brain to precisely locate seizure origins. For instance, the EMU at the Toronto Western Hospital annually manages about 300 patients for EEG recordings and nearly 40 patients for iEEG recordings.

Accurate detection and localization of seizure onset zones (SOZ) are crucial for effective treatment, particularly for those who are candidates for surgical intervention. Despite the significant volume of data generated, there is a lack of standardized methods for the automated processing, analysis, and interpretation of EEG and iEEG data, which hinders advancements in epilepsy treatment.

In recent years, machine-learning (ML) solutions have shown remarkable potential in automating tasks related to epilepsy management, such as seizure detection, characterization, prediction, and SOZ localization, with some works achieving accuracy levels exceeding 99% [4]–[16]. However, the full deployment of these technologies in clinical practice remains limited due to the lack of interpretability and transparency in ML models, often referred to as the “black-box” problem. Clinicians require models that not only provide high accuracy but also offer understandable and actionable insights to inform their decisions.

To address this challenge, research efforts have increasingly focused on developing explainability frameworks for ML models in epilepsy care [4]–[6], [17]–[19]. For example, Covert et. al. (2019) [4] explored localized and shared features across scalp EEG data to elucidate what their model was learning, linking this to the electrode influence in the model’s decision-making process. Pinto et. al. (2023) [5] provided a framework for explaining ML models, though further analysis was necessary to fully understand the brain dynamics of seizure generation. Batista et. al. (2024) [6] investigated preictal changes to predict seizures, emphasizing the need for models that clinicians can trust. Explainable AI (XAI) aims to make the decision-making processes of these models more transparent and interpretable, fostering greater trust and acceptance among healthcare professionals. Within this context, graph neural networks (GNNs) have emerged as a powerful paradigm, capable of effectively handling graph-structured data, such as the complex neural connections captured in iEEG recordings. For example, Lian et al. (2020) [15] introduced a joint graph structure and representation learning network, optimizing graph structure and preictal feature representations for seizure prediction. Their work highlights the potential of GNNs in providing interpretable and actionable insights for epilepsy management, bridging the gap between high-performance models and clinical application.

In this paper, we conduct an analysis of GNN models built for seizure detection from iEEG data, emphasizing model interpretability of spectral and spatial dependencies among iEEG channels. By utilizing two comprehensive iEEG datasets [20]–[22], comprising 23 and 16 subjects and 5 and 7 hours of iEEG recordings, we evaluate the performance of four different GNN architectures. Our results demonstrate that the highest-performing GNN model achieves a seizure detection accuracy of 97%, showcasing its potential for clinical application.

The contributions of this paper are twofold:

- We introduce a GNN-based framework for seizure detection from iEEG data that uses explainable and customizable data structures.
- We conduct cross-subject and cross-dataset experiments on four GNN models built for seizure detection.

By integrating advanced GNN techniques with explainable and customizable data structures, we aim to bridge the gap between high-performance ML models and their practical deployment in clinical environments, ultimately improving the management and treatment of epilepsy.

## II. Related Work

In this section, we review recent advancements in EEG signal processing using graph neural networks (GNNs) in conjunction with deep learning (DL) methods, particularly focusing on seizure detection and localization. Convolutional neural networks (CNNs) have traditionally been effective in learning from EEG data. However, their fixed-grid architecture limits their ability to capture complex electrode connections [23]–[27]. Recent studies have explored integrating GNNs with CNNs to enhance the representation of EEG data by leveraging graph structures [28].

Covert et al. (2019) [4] introduced a temporal graph convolutional network (GCN) model tailored for automated seizure detection from scalp EEG data. Their approach emphasized extracting localized and shared features across temporal sequences, demonstrating progress in understanding electrode influence during decision-making processes.

Hassan et al. (2019) [7] proposed an innovative method using feedforward neural networks (FfNNs) trained on multiband features derived from discrete wavelet transform (DWT) decomposition for epileptic seizure detection in EEG recordings. Meanwhile, Zeng et al. (2020) [14] developed a GCN model achieving near-perfect accuracy in seizure prediction from scalp EEG signals, although concerns about overfitting were raised.

Lian et al. (2020) [15] designed a joint graph structure and representation learning network aimed at optimizing graph structure and preictal feature representations for seizure prediction. Their focus on brain-computer interface (BCI)-aided neurostimulation systems highlighted the potential of integrating graph-based learning with clinical applications.

Zhao et al. (2021) [8] introduced a linear GCN approach to enhance feature embedding of raw EEG signals during seizure and non-seizure periods, reporting robust performance metrics including accuracy, specificity, sensitivity, F1, and AUC scores.

Dissanayake et al. (2021) [28] proposed a GNN model for seizure prediction using scalp EEG data, achieving state-of-the-art performance with over 95% accuracy. However, challenges in defining the prior graph for training highlighted ongoing methodological refinements needed in graph deep learning (GDL).

Li et al. (2022) [9] developed a graph-generative network model for dynamic discovery of brain functional connectivity using scalp EEG. Their supervised learning approach classified between ictal and non-ictal states with 91% accuracy, showcasing advancements in understanding brain dynamics through graph-based models.

Jia et al. (2022) [13] addressed the challenge of data volume in traditional DL models by employing a GCN model with 60-second windows to predict epileptic seizures from scalp EEG, aiming to make these technologies more suitable for wearable devices.

Liu et al. (2022) [29] combined unsupervised and semi-supervised GCN methods for SOZ localization using iEEG data, demonstrating improved precision compared to traditional indices like the Epileptogenicity Index (EI) [30] and Connectivity Epileptogenicity Index (cEI) [31].

Grattarola et al. (2022) [12] incorporated attention mechanisms into their GNN model for SOZ localization in epilepsy patients using iEEG recordings, identifying crucial brain regions associated with electrodes during interictal and ictal phases.

Wang et al. (2022) [11] developed a Spatiotemporal Graph Attention Network (STGAT) based on phase locking values (PLVs) to capture connectivity information among EEG channels, demonstrating high accuracy, specificity, and sensitivity in temporal and spatial learning. Additionally, Wang et al. (2023) [10] proposed a Weighted Neighbour Graph (WNG) representation for EEG signals, which aimed to reduce redundant edges by exploring different thresholding methodologies. Their study focused on improving the efficiency of graphbased EEG signal processing techniques.

Rahmani et al. (2023) [32] combined meta-learning with GNNs to offer personalized seizure detection and classification using minimal EEG data samples, achieving promising results in accuracy and F1-score.

Pinto et al. (2023) [5] reviewed explainability features in ML models for clinicians, emphasizing the need for deeper understanding of seizure dynamics and the interpretability of automated systems.

Statsenko et al. (2023) [16] evaluated a DL model for scalp EEG classification in binary and multi-class settings, exploring the influence of sampling rate and electrode number on model performance.

Raeisi et al. (2023) [33] integrated CNNs with graph attention networks (GATs) for EEG data from neonatal subjects, achieving high accuracy in seizure detection by capturing critical channel pairs and brain interareal information flow.

Batista et al. (2024) [6] investigated preictal changes in EEG data to predict seizures within 5 minutes of onset, highlighting challenges in real-life applications due to conservative alarm triggers.

Our study diverges from the aforementioned research by focusing on constructing effective and explainable graph representation (GR) data structures tailored to intracranial EEG (iEEG) data. Unlike previous works primarily centered on scalp EEG data, our emphasis on iEEG data offers superior resolution and volume, providing unprecedented insights into the intricate mechanisms underlying epilepsy. This strategic shift underscores the critical importance of leveraging iEEG data to advance our understanding and enhance clinical decision-making in epilepsy management. Moreover, we conduct an analysis of leveraging different GNN architectures.

## III. Methods and materials

### A. On the data

#### 1) OpenNeuro ds003029 dataset

We used a publicly available dataset hosted at OpenNeuro with Accession Number ds003029 [20], which was first used in Li et. al. (2021) [34], where the authors proposed the metric of neural fragility as a biomarker of epilepsy that can be used for SOZ localization. The dataset consists in iEEG and EEG data from 100 individuals across 5 epilepsy centers in the US that were resective surgery candidates. However, a large portion of the data was simply not available because one of the research centres failed to de-identify and share it. In the OpenNeuro repository online there are 35 data entries. However, for the purposes of our study, 12 entries were not usable due to inconsistencies in the clinical annotations. For example, in some cases there were not marks for the seizure onset or the seizure offset events, which posed a significant limitation given the supervised learning configuration of our GNN models. Thus, we only used 23 subjects from this dataset, which we describe in Table I. All subjects were monitored with electrocorticography (ECoG) electrodes. All subjects underwent resective surgery, except for UMMC001 and UMMC007. The surgery outcome was set as success or failure, where the former meant that no seizure events were registered on the patient after surgery, and the latter that seizure events continued to occur. The postsurgery monitoring period varied from patient to patient, with the shorter monitoring period being 1 year and the longest 7 years.

**TABLE I:**
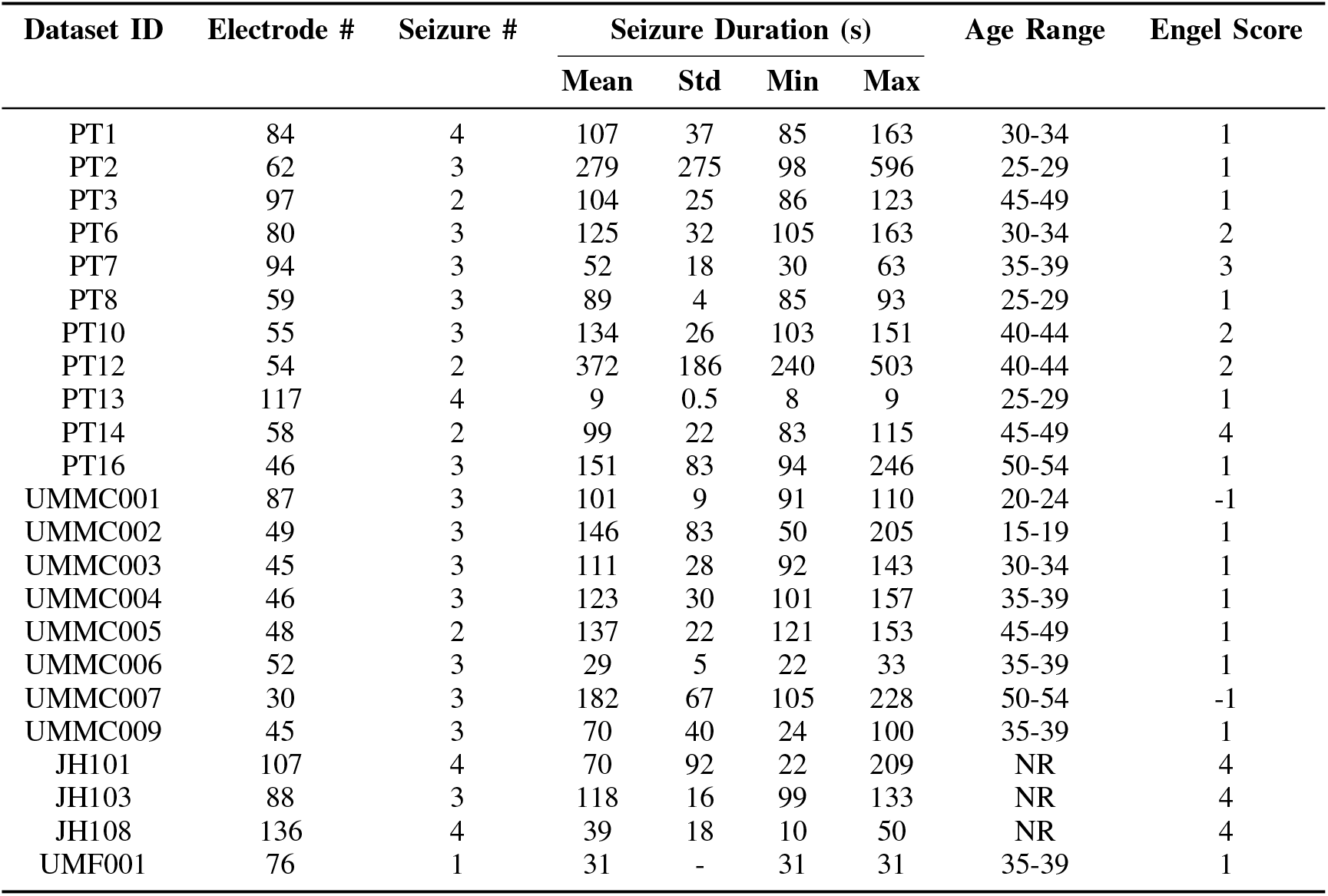
Summary of 23 patients’ information from the OpenNeuro ds003029 dataset. All data used for this study are already deidentified and publicly available online at OpenNeuro with Accession Number ds003029 [20]

| Dataset ID | Electrode # | Seizure # | Seizure Duration (s) |  |  |  | Age Range | Engel Score |
| --- | --- | --- | --- | --- | --- | --- | --- | --- |
|  |  |  | Mean | Std | Min | Max |  |  |
| PT1 | 84 | 4 | 107 | 37 | 85 | 163 | 30-34 | 1 |
| PT2 | 62 | 3 | 279 | 275 | 98 | 596 | 25-29 | 1 |
| PT3 | 97 | 2 | 104 | 25 | 86 | 123 | 45-49 | 1 |
| PT6 | 80 | 3 | 125 | 32 | 105 | 163 | 30-34 | 2 |
| PT7 | 94 | 3 | 52 | 18 | 30 | 63 | 35-39 | 3 |
| PT8 | 59 | 3 | 89 | 4 | 85 | 93 | 25-29 | 1 |
| PT10 | 55 | 3 | 134 | 26 | 103 | 151 | 40-44 | 2 |
| PT12 | 54 | 2 | 372 | 186 | 240 | 503 | 40-44 | 2 |
| PT13 | 117 | 4 | 9 | 0.5 | 8 | 9 | 25-29 | 1 |
| PT14 | 58 | 2 | 99 | 22 | 83 | 115 | 45-49 | 4 |
| PT16 | 46 | 3 | 151 | 83 | 94 | 246 | 50-54 | 1 |
| UMMC001 | 87 | 3 | 101 | 9 | 91 | 110 | 20-24 | -1 |
| UMMC002 | 49 | 3 | 146 | 83 | 50 | 205 | 15-19 | 1 |
| UMMC003 | 45 | 3 | 111 | 28 | 92 | 143 | 30-34 | 1 |
| UMMC004 | 46 | 3 | 123 | 30 | 101 | 157 | 35-39 | 1 |
| UMMC005 | 48 | 2 | 137 | 22 | 121 | 153 | 45-49 | 1 |
| UMMC006 | 52 | 3 | 29 | 5 | 22 | 33 | 35-39 | 1 |
| UMMC007 | 30 | 3 | 182 | 67 | 105 | 228 | 50-54 | -1 |
| UMMC009 | 45 | 3 | 70 | 40 | 24 | 100 | 35-39 | 1 |
| JH101 | 107 | 4 | 70 | 92 | 22 | 209 | NR | 4 |
| JH103 | 88 | 3 | 118 | 16 | 99 | 133 | NR | 4 |
| JH108 | 136 | 4 | 39 | 18 | 10 | 50 | NR | 4 |
| UMF001 | 76 | 1 | 31 | - | 31 | 31 | 35-39 | 1 |

Each patient dataset consists in 1 to 4 runs of seizure activity of varying duration. A run is considered as a single recording of iEEG activity that captures one seizure event, and is characterized by containing three time periods, the preictal, ictal, and postictal periods, which correspond to instances of time before, during, and after the seizure event. Each run that we selected has been clinically annotated for seizure onset and seizure offset times. The monitoring resolution across patients varied from 250 Hz, 500 Hz, and 1 kHz. We have selected 67 runs in total for the 23 patients, with an average number of electrodes of 71.86 ± 26.32, the average number of seizures in the dataset is 2.86 ± 0.66, and the average seizure duration time in seconds is 118.91 ± 78.82.

#### 2) SWEC-ETHZ short-term dataset

We used another publicly available dataset hosted by the Sleep-Wake-Epilepsy-Center (SWEC) of the University Department of Neurology at the Inselspital Bern and the Integrated Systems Laboratory of the ETH Zurich at http://ieeg-swez.ethz.ch/. This was first used by Burrello et. al. (2018 and 2019) [21], [22], where they investigated the problem of seizure detection using hyperdimensional computing. The dataset consists in 100 anonymized intracranially recorded electroencephalographic (iEEG) datasets of 16 patients with pharmaco-resistant epilepsy who were evaluated for epilepsy surgery.

The following description was extracted from the online repository on May 2024: *the iEEG signals were recorded intracranially by strip, grid, and depth electrodes. After 16-bit analog-to-digital conversion, the data were digitally bandpass filtered between 0*.*5 and 150 Hz using a fourth-order Butterworth filter prior to analysis and written onto disk at a rate of 512 Hz. Forward and backward filtering was applied to minimize phase distortions. All the iEEG recordings were visually inspected by an EEG board-certified experienced epileptologist (K*.*S*.*) for identification of seizure onsets and endings and exclusion of channels continuously corrupted by artifacts. Each recording consists of 3 minutes of preictal segments (i*.*e*., *immediately before the seizure onset), the ictal segment (ranging from 10 s to 1002 s), and 3 minutes of postictal time (i*.*e*., *immediately after seizure ending)*. For consistency with the OpenNeuro dataset, we only kept up to 4 seizure files per patient, though more seizure files are available in a per-patient basis. A summary of the data is shown in Table II, which was adapted from [22] and modified to the number of seizures and seizure duration statistics. We have selected 55 runs in total for the 16 patients, with an average number of electrodes of 61.93 ± 18.77, the average number of seizures in the dataset is 3.37 ± 0.74, and the average seizure duration time in seconds is 116.25 ± 107.27.

**TABLE II:** Summary of 16 patients’ information from the SWEC-ETHZ dataset. All data used for this study are already deidentified and publicly available online at http://ieeg-swez.ethz.ch/

| Dataset ID | Electrode # | Seizure # | Seizure Duration (s) |  |  |  | Age Range | Engel Score |
| --- | --- | --- | --- | --- | --- | --- | --- | --- |
|  |  |  | Mean | Std | Min | Max |  |  |
| 1 | 47 | 4 | 85 | 57 | 10 | 136 | 20-24 | 2 |
| 2 | 42 | 4 | 223 | 88 | 96 | 301 | 15-19 | 1 |
| 3 | 98 | 2 | 99 | 36 | 73 | 125 | 25-29 | 1 |
| 4 | 62 | 4 | 140 | 14 | 127 | 160 | 30-34 | 4 |
| 5 | 54 | 4 | 105 | 33 | 84 | 154 | 20-24 | 2 |
| 6 | 64 | 4 | 145 | 47 | 89 | 190 | 45-49 | 1 |
| 7 | 36 | 2 | 15 | 1 | 14 | 16 | 30-34 | 4 |
| 8 | 59 | 2 | 56 | 6 | 52 | 61 | 35-39 | 4 |
| 9 | 56 | 4 | 119 | 11 | 104 | 129 | 25-29 | 1 |
| 10 | 100 | 4 | 12 | 1 | 10 | 14 | 45-49 | 2 |
| 11 | 64 | 2 | 109 | 36 | 83 | 135 | 25-29 | 1 |
| 12 | 49 | 4 | 36 | 10 | 23 | 46 | 55-59 | 4 |
| 13 | 92 | 4 | 72 | 36 | 19 | 100 | 45-49 | 2 |
| 14 | 74 | 4 | 493 | 313 | 154 | 903 | 35-39 | 1 |
| 15 | 61 | 3 | 120 | 66 | 52 | 184 | 20-24 | 4 |
| 16 | 59 | 4 | 80 | 9 | 67 | 89 | 30-34 | 2 |

### B. Data processing pipeline

The data processing pipeline that we developed is illustrated in Figure 1. It is entirely developed in Python, and it uses the MNE software [35] for iEEG data management and preprocessing, the Spektral project [36] and Keras [37] with Tensorflow [38] for data balancing and GNN model handling, and it leverages the supercomputer infrastructure from the Canada-wide High Performance Computing platform from the Digital Research Alliance of Canada. The technical integration of these tools with our customized software interfaces will be disseminated elsewhere.

**Fig. 1:**
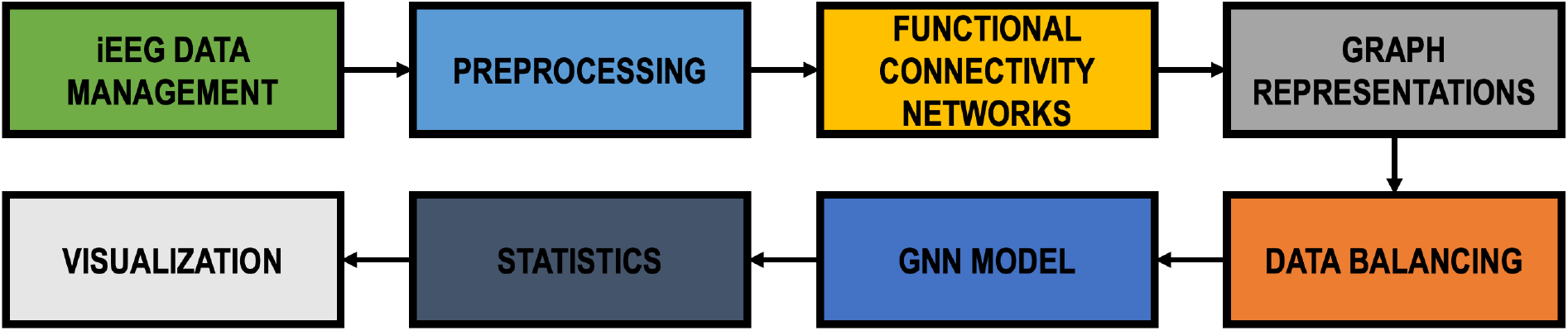
Diagram illustrating our data pipeline. The tool is entirely developed in Python, and it is powered by MNE [35] for iEEG data management and preprocessing, Spektral [36] and Keras [37] with Tensorflow [38] for data balancing and GNN model handling, and it is running on the Canada wide High Performance Computing platform managed by the Digital Research Alliance of Canada.

#### 1) iEEG data management

Monitoring an epilepsy patient with iEEG for a 24/7 period collects around 10 TB (terabytes) of data. Handling iEEG data is a concern of every hospital or EMU, and it is influenced by the monitoring equipment available to these entities, and the clinical and technical expertise of the individuals working with the data. This unstandardized practice has resulted in large heterogeneous iEEG datasets that are not findable, accessible, interoperable and reusable (FAIR), and that have limited clinical use within their own centers. Thus, iEEG data management standardization protocols are required. In 2019, the iEEG-BIDS protocol was proposed in [39], which extends the Brain Imaging Data Structure (BIDS) protocol [40] to operate with iEEG data. Since then, the iEEG-BIDS protocol has been spread around the iEEG research and clinical community, although its adoption as a gold-standard has yet to be realized. The OpenNeuro ds003029 dataset [20] is stored under the iEEG-BIDS format. Within our pipeline, we leverage iEEG-BIDS data handling processes with the MNE software [41], which provides robust tools to read/write and process this type of data. In contrast, the SWEC-ETHZ dataset is stored in a customized data format leveraging Python NumPy arrays.

#### 2) Data preprocessing

Although iEEG data can be captured at high sampling rates of up to 25KHz [42], clinical practice often relies on lower recording resolutions from 250Hz to 2kHz [43]. Despite the availability of high resolution sensors, the monitoring process is far from perfect, and recordings are often corrupted due to many-form noise sources. These include power-line electricity noise and artifacts caused by involuntary body movements (i.e., eye-lid or muscle movement). Moreover, it is common to have unusable data from bad channels due to bad electrode placement or contact. Data corruption phenomena are commonly fixed by preprocessing the data, which is yet another unstandardized process in clinical and research practice.

Our iEEG data preprocessing pipeline consists in first removing bad channels from the dataset which were identified by the clinicians, and are part of the metadata found within the iEEG-BIDS format in the OpenNeuro dataset. The SWEC-ETHZ dataset already contains only good channels. Next we apply a notch filter at 60 Hz and corresponding harmonics (120 Hz, 180 Hz, etc.), to attenuate the presence of powerline noise. We do not perform any artifact removal or artifact reconstruction method as we are interested in using the data with as little preprocessing as possible. This part of our pipeline also leverages the MNE software tools for iEEG signal processing.

The electrophysiological activity of the brain is known to use different frequency bands for different purposes. For instance, recently low-gamma oscillations have been found to be involved in emotional regulation, and high-gamma oscillations seem to be involved in speech temporal information [44], [45]. However, the self-regulatory neuromodulation processes of the brain are vastly unknown. Consequently, we are also interested in investigating the role of different frequency bands and their role in seizure events. Moreover, we are interested in devising how to use this information to create powerful iEEG-GRs to improve seizure detection with GNNs. For this, our preprocessing pipeline also includes bandpass filtering with a highpass filter set at 0.1 Hz and a lowpass filter set at the Nyquist limit, which depends on the sampling frequency of each run. Last, we clip each preprocessed run in their preictal, ictal, and postictal signal traces, and stored them in serialized file objects.

#### 3) Functional connectivity networks

FCNs are abstractions of brain data that aim to represent the dynamics of neurophysiological activity recorded with neuroimaging tools, such as diffusion tensor imaging (DTI) or electrophysiological tools, such as iEEG. FCNs are useful to study neurophysiological activity from a networks perspective, and they can be used to map network-modeled neurophysiological activity to behavioral and cognitive dimensions. Network analysis of iEEG data pose new perspectives to uncover neural circuits underlying neurological disorders, which will be instrumental for the development of new treatment options [46].

In this study, we focus solely on iEEG-based FCNs. Thus, the purpose of the iEEG data-to-network abstraction is to quantify the degree of similarity across iEEG signals. While there are several methods to create iEEG-based FCNs, mapping FCNs to behavioral or cognitive tasks is an active research field, and there is no one method that is useful for all cases. For this study, we consider the methods of Pearson correlation, which is used to measure the similarity of energy levels across signals over time, in the time domain; the coherence, which also measures similarity of energy levels across signals over time but in the frequency domain; and the phase-lock value (PLV), which measures where are the signals over time.

Our method for FCN creation is illustrated in Figure 2. In block **A**, we show an iEEG run from one patient, where the y-axis depicts the signals collected at each electrode (measuring energy in Volts), and the x-axis depicts time starting at *t*0 and ending at *L* seconds. For the binary classification problem we label nonictal data (preictal and postictal traces) as class 0, and ictal data as class 1. For the multi-class classification problem we label preictal, ictal, and postictal data as class 0, 1, and 2, respectively. There are two markers on each run, *t_on* and *t_off*, indicating the beginning and the end of the ictal activity as annotated by the clinical experts. To compute FCNs, we define a window, *W*, depicted in green at the top-left, which indicates the interval of time in which the degree of connectivity across signals is assessed. Then, we define a sliding window, *SW*, which indicates how to slide *W* across the run to create FCN sequences as depicted in blocks **B** and **C**. For this study, we consider 1 second *W* and 0.125 seconds *SW* .

**Fig. 2:**
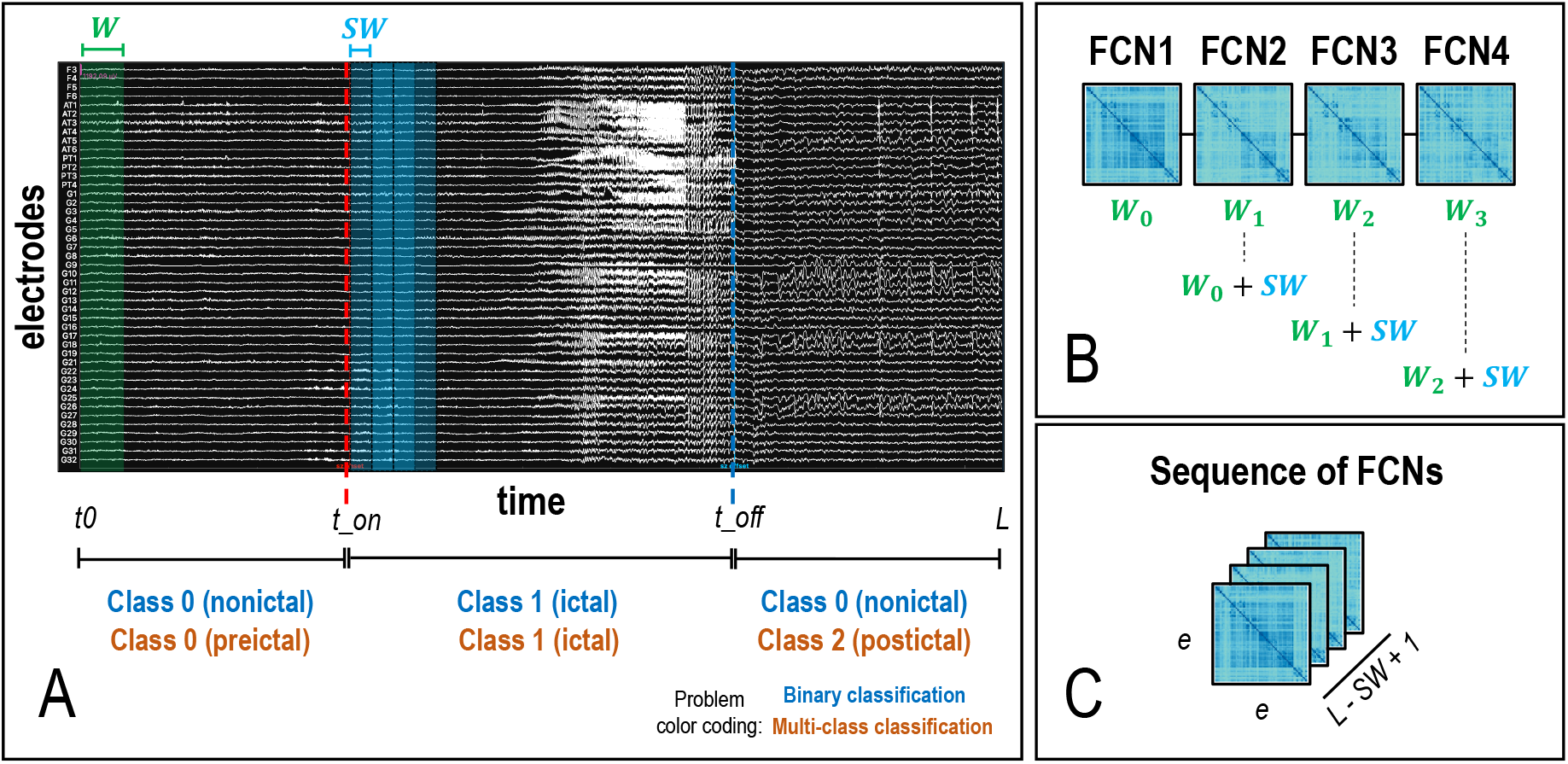
Methodology to create functional connectivity networks (FCNs). **A**. Every iEEG record (each run) of duration *L* has 2 main time marks, the seizure onset (*t_on*) and the seizure offset (*t_off*). The signal trace within *t*0 and *t on* indicates the preictal period of the signal, which we label as 0 for the binary and multi-class classification problems. The signal trace within *t_on* and *t_off* indicates the ictal period, which we label as 1 for the binary and multi-class classification problems. The signal trace within *t_off* and *L* indicates the postictal period, which we label as 0 and 2 for the binary and multi-class classification problems, respectively. To create FCNs, we declare a window *W* (in green, top-left) that indicates the portion of the iEEG record to analyze. Then, we declare a sliding window *SW* (in blue, top-center) that indicates how to slide *W* over the entire record. **B**. Illustration of 4 FCNs sequentially created by sliding *W* by *SW*. **C**. The result is a multidimensional array shaped by (*e, e, L* − *SW* + 1), representing an FCN sequence.

#### 4) Graph representations

In computer science, GRs are data structures that extend the original graph data structure to account for node and edge feature vectors. Take for instance a regular graph data structure illustrated in Figure 3, **A**, where *G* = {*V, E* }, and *V* and *E* are sets of vertices and edges between vertices. Instead, the *GR* in **B** is represented as 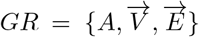, where *A* is an adjacency matrix or “original graph”, 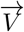 and 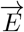 are sets of multidimensional vectors representing node and edge features [47]. As shown in block **C**, similarly to the creation of sequences of FCNs, we can create sequences of GRs to increment the representational power of the abstracted data.

**Fig. 3:**
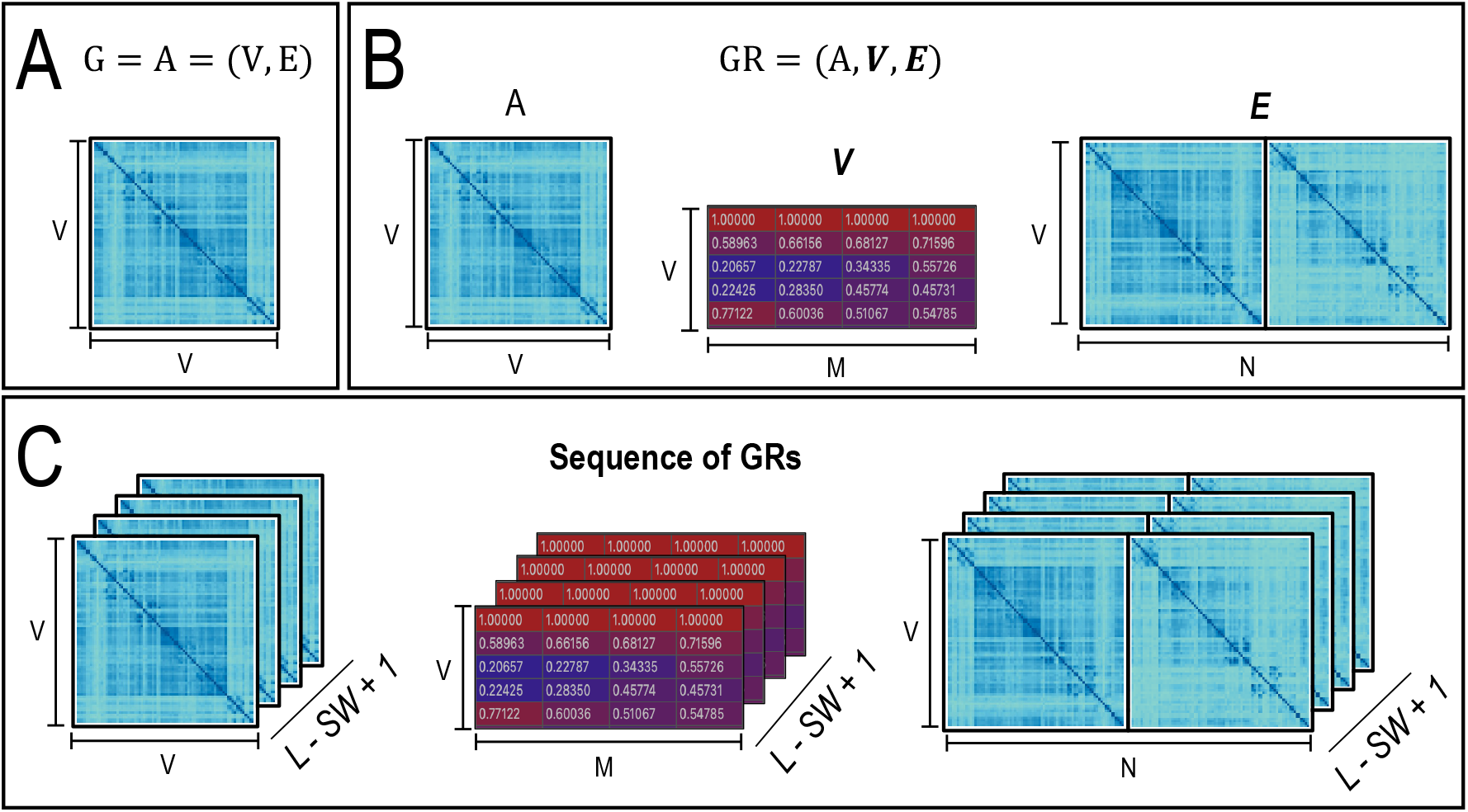
Illustration of graph representations (GRs). **A**. *G* = *A* = (*V, E*) is the original graph represented as a set of vertices *V* and edges *E*. **B**. *GR* = (*A*, **V, E**) is a graph representation with 3 elements: and adjacency matrix, *A*^*V* × *V*^, a node features vector, *V* ^*V* × *M*^, and an edge features vector, *E*^*V* × *N*^, where *M* and *N* are the number of node and edge features, respectively. **C**. Similar to the FCN sequences, we can create sequences of GRs.

#### 5) Data balancing

To evaluate the GNN model, we split each patient dataset in train, validation, and test sets, taken from 80%, 10%, 10% of the data, respectively. However, the amount of data samples varies from the preictal, ictal, and postictal signal traces, meaning that there is an imbalanced class representation for the binary and multi-class classification problems. Consequently, we implement a data balancing algorithm that guarantees there is a balanced representation of classes. For this, we consider the maximum number of ictal samples within a run to be the total number of ictal samples, and non-ictal samples are taken from the preictal and postictal traces in equal amounts. This process is illustrated in Figure 4. Each data sample represents a GR that is used for the train, validation, and test datasets, marked with a red cross, a cyan square, and a turquoise circle, respectively.

**Fig. 4:**
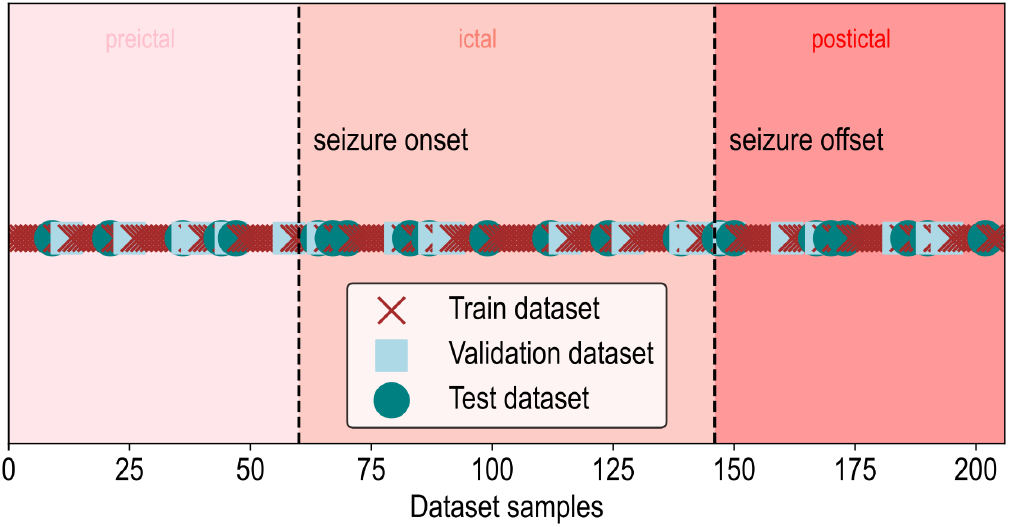
Illustration of the data balancing procedure.

### C. Graph neural network architecture

Our GNN architecture is depicted in Figure 5

**Fig. 5:**
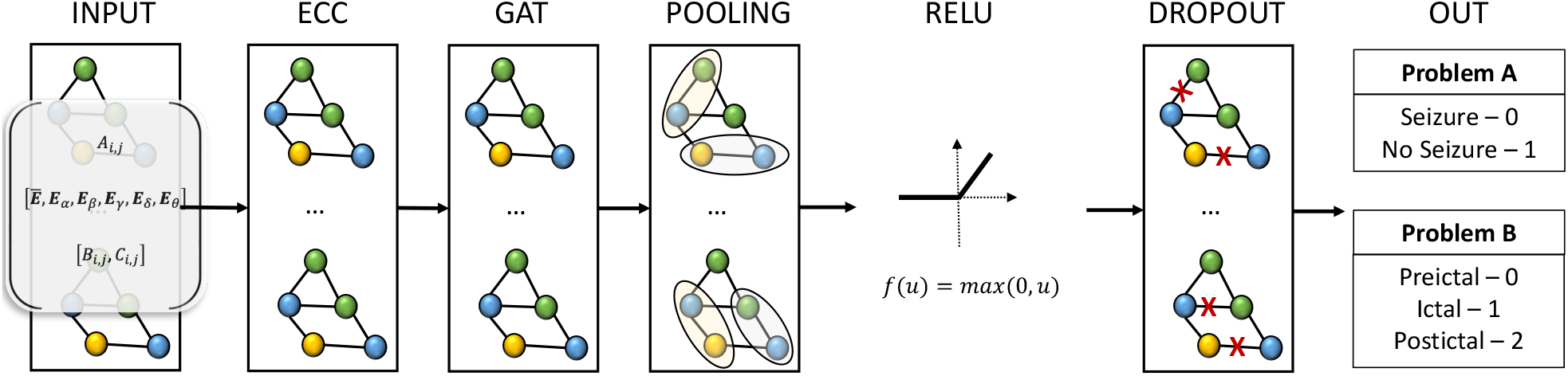
Architecture of the GNN model proposed by Grattarola et. at. in [12], extended for multi-class classification.

#### 1) ECC, edge-conditioned convolution layer

The GNN model proposed by Grattarola et. at. in [12] is illustrated in Figure 5. This depicts a 2-layer neural network composed of an ECC layer [48] followed by a GAT layer [49]. The ECC layer constrains the learning to focus on the edge features, while the GAT layer computes attention coefficients, which are measurements that quantify the importance of each node in a graph in the learning process. In [12], the authors proposed the use of the attention mechanism as a means to map attention coefficients to the SOZ. However, we did not find success in applying this methodology, and we plan to devise strategies for SOZ localization within our pipeline in future work.

The ECC layer as defined in Equation 1, serves as a critical component in processing the graph representation of EEG signals. In this equation, *Y* (*i*) represents the transformed feature at node *i*, obtained by aggregating information from neighboring nodes *j* within the graph *N* (*i*).

Thus, the ECC layer employs edge-conditioned convolutions to capture intricate relationships between nodes and edges, while the GAT layer leverages attention mechanisms to dynamically weight the contributions of neighboring nodes. These mathematical formulations constitute essential components of graph neural networks applied to the classification of EEG signals, facilitating the effective learning and representation of complex data patterns.

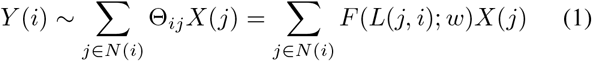

#### 2) GCN, graph convolutional layer

This layer computes:

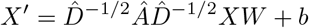

where *Â* = *A*+*I* is the adjacency matrix with added self-loops and 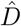 is its degree matrix.

#### 3) Diff, diffusion convolutional layer

Given a number of diffusion steps *K* and a row-normalized adjacency matrix *Â*, this layer calculates the *q*-th channel as:

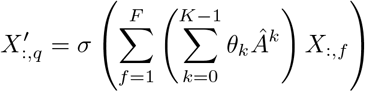

#### 4) Cheb, Chebyshev convolutional layer

This layer computes:

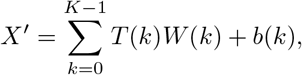

where *T* (0), …, *T* (*K* 1) are Chebyshev polynomials of 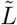 defined as:

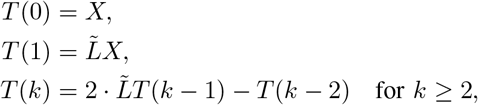

where

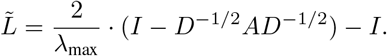

#### 5) GAT, graph attention layer

Conversely, the GAT layer introduces the concept of attention coefficients (*e*_*i*,*j*_) to assign varying degrees of importance to neighboring nodes during the aggregation process. In this equation, *a* signifies an attention mechanism that computes these coefficients based on the node features **h**_**i**_ and **h**_**j**_ associated with nodes *i* and *j*. The weight matrices **W** enable the model to adaptively learn the optimal attention weights.

This layer computes a convolution similar to `layers.GraphConv`, but uses the attention mechanism to weight the adjacency matrix instead of using the normalized Laplacian:

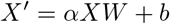

where

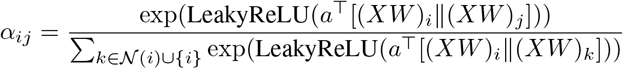

where *a* ∈ ℝ^2*F′*^ is a trainable attention kernel. Dropout is also applied to *α* before computing *Z*. Parallel attention heads are computed in parallel and their results are aggregated by concatenation or averaging.

#### 6) Pooling layer

The pooling or aggregation process is governed by the learned parameters Θ_*ij*_, which determine the significance of each neighboring node’s contribution. The function *F* (*L*(*j, i*); *w*) denotes a convolution operation applied to the edge feature *L*(*j, i*), with respect to a set of trainable weights *w*. This operation effectively combines information from neighboring nodes and their associated edge features, enabling the model to capture latent patterns and dependencies inherent in the graph structure.

### D. Code Availability

The entire graph representation and graph neural network pipeline has been containerized into a Python package, available at https://github.com/yousifKashef/NeurologyKit. To use the repository, sample scripts are located in the ‘projects’ directory and the Python modules are located under ‘bgreg’. Input data can be inserted under ‘data files’, enabling dataset customization.

## IV. Results

### A. Graph representations

#### 1) Leveraging node and edge features

Graph-structured data contained within GRs are commonly used as input data to build GNN models, a paradigm of artificial neural networks optimized to operate with graph-structured data. In recent years, GNN models have been used to advance the understanding of real-life network phenomena found across several scientific fields including physics and chemistry [47]. More recently, some works have started to show the potential of GNNs to assist the field of neuroscience [12], [50], but further research is require to deploy these models in the clinical practice. Importantly, the ability of GNN models to learn about the graphs they operate on is dependant on the abstraction of the data within the GRs, which is still considered an art in the realm of deep learning and AI. In this study, we investigate the creation of GRs of iEEG data that are more helpful to build a GNN model for seizure detection.

Our goal is to understand what combination of graph representation elements renders the most powerful iEEG-GR for seizure detection. We consider 9 GRs, presented in Table III, which we cluster in 3 groups. Group 1 is composed of tests b1, b2, and b3 correspond to the baseline GRs used in [12] (b1 and b2), where node and edge features are considered as all-ones vectors, and adjacency matrices are considered as FCNs of the Pearson correlation and PLV methods. Test b3 was not used in [12], but we include it to evaluate the usage of the coherence method in this form. Group 2 is composed of tests t11, t12, and t13, and correspond to GRs that use the average energy recorded by the iEEG electrodes as node features vector and FCNs of the Pearson correlation, coherence, and PLV methods. The average energy at electrode vector has a shape of (*V*, 1), where V is the number of electrodes. Last, group 3 are tests t21, t22, and t23 and correspond to GRs that consider the average energy at electrode and the average energy at electrode by frequency band as node features vector. The average energy at electrode by frequency band has a shape of (*V, F*), where *F* corresponds to the frequency bands considered. For iEEG signals that were recorded with a 250 Hz sampling rate, *F* = 6, as we consider the frequency bands *δ*, delta (1 - 4 Hz), *θ*, theta (4 - 8 Hz), *α* (8 - 13 Hz), alpha *β*, (13 - 30 Hz), *γ*, gamma (30 - 70 Hz), and Γ, high-gamma (70 - 100 Hz). For signals that were recorded with a 500 Hz sampling rate, *F* = 7, as we also consider ripples in the frequency band (100 – 250 Hz). For signals recorded at 1 kHz sampling rate, *F* = 8, as we also consider fast ripples in the frequency band (250 - 500 Hz). We then create the node features vectors for these tests by concatenating the vectors for the average energy at electrode and the average energy at electrode by frequency band. Similarly, for the edge features vector we concatenate 2 FCNs that are different from the adjacency matrix. For example, in test t21, the adjacency matrix is considered to be a Pearson correlation-based FCN, while the edge features vector is the concatenation of the coherence FCN with the PLV FCN.

**TABLE III:** Graph representation elements.

| Test | AM | Node Features | Edge Features |
| --- | --- | --- | --- |
| b1 | PC | **1** | **1** |
| b2 | COH | **1** | **1** |
| b3 | PLV | **1** | **1** |
| t11 | PC | $E(e)$ | COH |
| t12 | PC | $E(e)$ | PLV |
| t21 | COH | $E(e)$ | PC |
| t22 | COH | $E(e)$ | PLV |
| t31 | PLV | $E(e)$ | PC |
| t32 | PLV | $E(e)$ | COH |

AM: Adjacency matrix metric and window time, GNN: Graph neural network layers, PC: Pearson correlation, PLV: phase-lock value, Coh: coherence, Coh+: average overall coherence and coherence by frequency band ( $\lambda$ ) **1**: all-ones vector, $E(e)$ : average energy at electrode ( $e$ ), $E(e, \lambda)$ : average energy at electrode ( $e$ ) by frequency band ( $\lambda$ ).

Table IV shows the results of binary classification. Note that all tests with non-trivial node and edge features (t11-t32) outperform the baseline tests with all-ones vector node and edge features (b1-3).

**TABLE IV:**
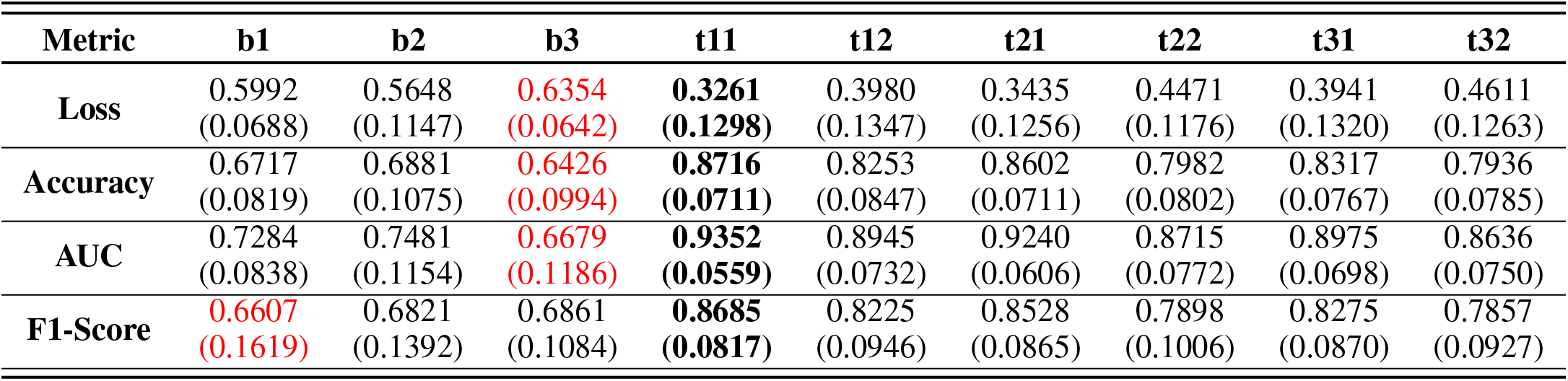
Mean (SD) performance evaluation of GNN model for binary classification across 23 subjects.

| Metric | b1 | b2 | b3 | t11 | t12 | t21 | t22 | t31 | t32 |
| --- | --- | --- | --- | --- | --- | --- | --- | --- | --- |
| Loss | 0.5992<br>(0.0688) | 0.5648<br>(0.1147) | 0.6354<br>(0.0642) | 0.3261<br>(0.1298) | 0.3980<br>(0.1347) | 0.3435<br>(0.1256) | 0.4471<br>(0.1176) | 0.3941<br>(0.1320) | 0.4611<br>(0.1263) |
| Accuracy | 0.6717<br>(0.0819) | 0.6881<br>(0.1075) | 0.6426<br>(0.0994) | 0.8716<br>(0.0711) | 0.8253<br>(0.0847) | 0.8602<br>(0.0711) | 0.7982<br>(0.0802) | 0.8317<br>(0.0767) | 0.7936<br>(0.0785) |
| AUC | 0.7284<br>(0.0838) | 0.7481<br>(0.1154) | 0.6679<br>(0.1186) | 0.9352<br>(0.0559) | 0.8945<br>(0.0732) | 0.9240<br>(0.0606) | 0.8715<br>(0.0772) | 0.8975<br>(0.0698) | 0.8636<br>(0.0750) |
| F1-Score | 0.6607<br>(0.1619) | 0.6821<br>(0.1392) | 0.6861<br>(0.1084) | 0.8685<br>(0.0817) | 0.8225<br>(0.0946) | 0.8528<br>(0.0865) | 0.7898<br>(0.1006) | 0.8275<br>(0.0870) | 0.7857<br>(0.0927) |

In Table V we show the evaluation of the GNN model using the same GRs from the binary classification problem for the multi-class classification problem. The base tests (b1-3) are excluded as their relatively lower performance had already been indicated through the binary problem. Here, we also see that by including more data within the GRs we can significantly improve the performance of the GNN model in accuracy, AUC, and F1-score with mean performance across the 25 subjects of 90%, 97%, and 88% for tests t21, t22, and t23. Importantly, we can see that the GNN model is capable of discriminating between preictal and postictal signal traces, showing a potential to develop seizure prediction algorithms in the future.

**TABLE V:** Mean (SD) performance evaluation of GNN model for multi-class classification across 23 subjects.

| Metric | t11 | t12 | t21 | t22 | t31 | t32 |
| --- | --- | --- | --- | --- | --- | --- |
| Loss | 0.4050<br>(0.1881) | 0.5047<br>(0.2078) | 0.4254<br>(0.1882) | 0.5809<br>(0.1973) | 0.5073<br>(0.2042) | 0.5885<br>(0.1844) |
| Accuracy | 0.8504<br>(0.0892) | 0.7961<br>(0.1101) | 0.8369<br>(0.0933) | 0.7649<br>(0.1055) | 0.7975<br>(0.1047) | 0.7615<br>(0.0991) |
| AUC | 0.9545<br>(0.0440) | 0.9240<br>(0.0643) | 0.9479<br>(0.0467) | 0.9082<br>(0.0655) | 0.9241<br>(0.0640) | 0.9065<br>(0.0639) |
| F1-Score | 0.8241<br>(0.1051) | 0.7597<br>(0.1295) | 0.8105<br>(0.1059) | 0.7260<br>(0.1211) | 0.7633<br>(0.1254) | 0.7195<br>(0.1186) |

In general, our methodology is better than the baseline, however, same GR structures do not work equally well for all patients, as depicted by the outliers in these figures. We speculate that similarly to the difficulties encountered by clinical experts in visually inspecting the iEEG data, GNN models struggle to learn features from ambiguous or corrupted data recordings. In future work, we will investigate the relationships between graph embeddings and iEEG signal characteristics, and their impact on GNN learning and decision-making processes.

It’s worth noting that our methodology is constrained by the features utilized for GR creation, as detailed in Table III. The three distinct methods we have employed for computing FCNs from iEEG data serve as gateways to different data abstractions. Each method offers a unique lens through which we can decipher the underlying intricacies of the data.

Firstly, the Pearson correlation method allows us to quantify the degree of similarity in signal activity based on the energy levels observed at any given moment in time. This approach, with its focus on the temporal energy perspective, is particularly valuable when scrutinizing iEEG patterns within the time domain. It sheds light on how signal energies evolve over time, enabling us to discern subtle variations and transitions in the data.

Secondly, coherence analysis steps into the frequency domain, providing us with a distinct perspective on energy levels within iEEG data. By quantifying energy levels in the frequency domain, coherence analysis facilitates the study of iEEG patterns from a frequency-based viewpoint. This methodology unveils how different frequency components interact and synchronize, offering a richer understanding of the data’s dynamics.

Thirdly, PLV analysis offers a nuanced dimension by quantifying the degree of synchronicity among signals over time. It enables us to delve deep into the temporal dynamics of iEEG data, elucidating how signals align and interact temporally. Thus, PLV helps to understand signal synchronization.

To explore further possibilities and delve into higher-level processes, such as directional signaling with the direct absolute coherence (DAC) [51], we anticipate undertaking future investigations. This broader exploration will help us uncover new insights and refine our approach.

#### 2) Different adjacency matrix lengths

We aim to establish an optimal time span for the window used in the computation of the adjacency matrix metric. Given its superior performance as illustrated in the previous section, we use coherence as the adjacency metric in our tests. In Figure 6, we show that increases in adjacency matrix window time span from 1 to 20 seconds trends toward greater performance in test accuracy, F1-Score, and AUC. Moreover, the 20s time span shows statistically significantly better performance in all aforementioned metrics among the patient dataset compared to all the time spans 5s and shorter. (*p* ≤ 0.05 from Man-Whitney U test). There are no sacrifices in training time, as represented in Figure 6d. Note that attempting to compute adjacency matrices for time spans beyond 20s resulted in computation problems, such as memory overflow issues, largely due to artefacts and errors present in the original dataset. Thus, for our purposes we conclude that 20s is the optimal time span for computation of the adjacency matrix in GRs in order to maximize performance.

**Fig. 6:**
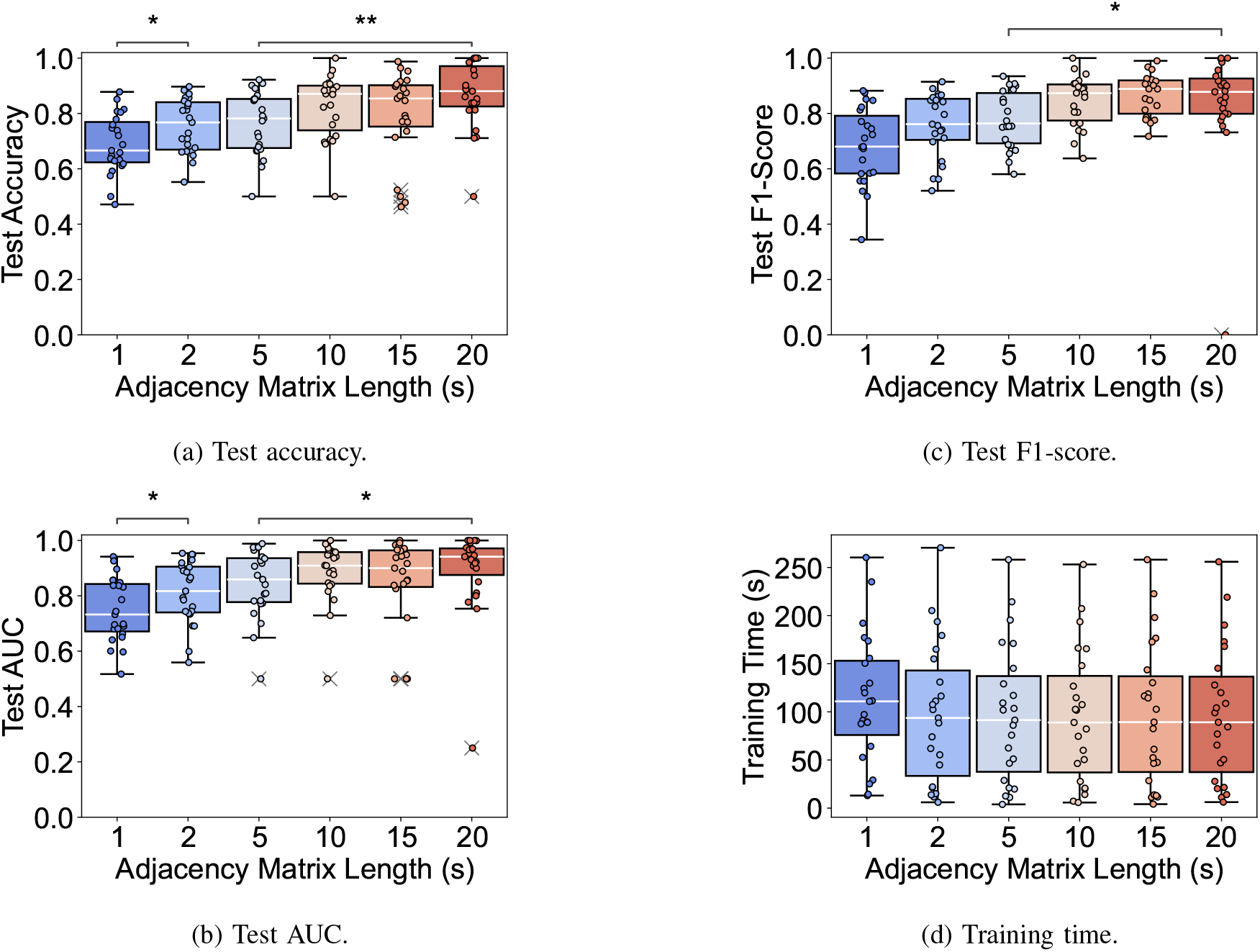
Test performance and training time for binary classification of seizure detection from graph representations with varying window times for their adjacency matrix computation. Coherence is used as the adjacency matrix metric. A Man-Whitney U test is run (* indicates *p* ≤ 0.05, ** indicates *p* ≤ 0.01).

#### 3) Extending node and edge features

We first explore the effects of increasing the information stored in GR node features by expanding their dimensions. Node features are expanded to include both average overall energy as well as average energy by frequency band, potentially yielding up to 9 total dimensions depending on on the original data. We use phase locking value as edge features and 20s windows of coherence as the adjacency matrix metric. Using the multi-task classification problem, we obtain mean loss (SD) 0.1741 (0.1126), mean accuracy (SD) 95.26% (3.57%), mean AUC (SD) 0.9906 (0.0115), and mean F1-Score (SD) 0.9411 (0.0441). This demonstrates improvement over all tests t11-32 from Table V.

Next we explore expanding edge features. We use the same setup as above (including the expanded node features), except now edge features are expanded to include both average overall coherence as well as average coherence by frequency band. Again, we see a noticeable improvement in all performance metrics: mean loss (SD) 0.1504 (0.2548), mean accuracy (SD) 97.07% (2.62%), mean AUC (SD) 0.9942 (0.0065), and mean F1-Score (SD) 0.9616 (0.0335).

### B. GNN models

We will now vary the first layer of our GNN model, while keeping the second layer constant as a graph attention layer (GAT). To demonstrate robustness of our GR methodology and the GNN models, we conduct tests on 2 separate open-source datasets: OpenNeuro (Table VI), which was used in all previous testing, and SWEC-ETHZ (Table VII). Both datasets yield similar results, reflecting the stability of our GR pipeline. For both datasets, the edge conditioned convolutional (ECC) layer performed strongest while the diffusion convolution layer was weakest.

**TABLE VI:** Mean (SD) performance evaluation of different GNN Layers for multi-class classification across OpenNeuro dataset.

| Metric | ChebGAT | DiffGAT | ECCGAT | GCNGAT |
| --- | --- | --- | --- | --- |
| **Loss** | 0.3377<br>(0.1486) | **0.5286**<br>**(0.3162)** | **0.1554**<br>**(0.3205)** | 0.2383<br>(0.1158) |
| **Accuracy** | 0.9023<br>(0.0618) | **0.7788**<br>**(0.1528)** | **0.9703**<br>**(0.0273)** | 0.9353<br>(0.0417) |
| **AUC** | 0.9757<br>(0.0233) | **0.8996**<br>**(0.1171)** | **0.9943**<br>**(0.0069)** | 0.9868<br>(0.0116) |
| **F1-Score** | 0.8831<br>(0.0752) | **0.6865**<br>**(0.2401)** | **0.9611**<br>**(0.0353)** | 0.9200<br>(0.0503) |

**TABLE VII:** Mean (SD) performance evaluation of different GNN Layers for multi-class classification across ETHZ dataset.

| Metric | ChebGAT | DiffGAT | ECCGAT | GCNGAT |
| --- | --- | --- | --- | --- |
| **Loss** | 0.3115<br>(0.1749) | **0.7174**<br>**(0.2987)** | **0.1327**<br>**(0.1564)** | 0.2097<br>(0.1412) |
| **Accuracy** | 0.9135<br>(0.0873) | **0.6858**<br>**(0.1529)** | **0.9621**<br>**(0.0674)** | 0.9452<br>(0.0619) |
| **AUC** | 0.9756<br>(0.0433) | **0.8346**<br>**(0.1259)** | **0.9897**<br>**(0.0268)** | 0.9876<br>(0.0237) |
| **F1-Score** | 0.8945<br>(0.1120) | **0.5630**<br>**(0.2567)** | **0.9517**<br>**(0.0851)** | 0.9333<br>(0.0760) |

## V. Conclusion

Automating the process of iEEG-based seizure detection within clinical pipelines treating epilepsy is a timely need to improve healthcare practice. In this paper we proposed an iEEG data processing pipeline that uses a GNN model to detect ictal and non-ictal activity, and that detects preictal, ictal, and postictal data. Moreover, we demonstrate that by leveraging iEEG signal data as GRs of iEEG data we can significantly improve the performance of GNN-based systems.

## Data Availability

All data produced are available online at: 1) The OpenNeuro Epilepsy-iEEG-Multicenter-Dataset with accession number ds003029, accessible through https://openneuro.org/datasets/ds003029/versions/1.0.7; 2) The SWEC-ETHZ iEEG Database, accessible through http://ieeg-swez.ethz.ch/.

https://openneuro.org/datasets/ds003029/versions/1.0.7

## Acknowledgements

We thank the epilepsy monitoring unit at the Toronto Western Hospital in Toronto, Canada for sharing their experience in dealing with epilepsy patients and in explaining the challenges in bridging data scientists to epilepsy care. We also thank Professors Taufik Valiante and Mary Pat McAndrews at the Krembil Research Institute for their valuable input and suppor in the conceptualization of this project. As well, we thank Dr. Daniele Grattarola for sharing his technical expertise in the development of graph neural network models.

---

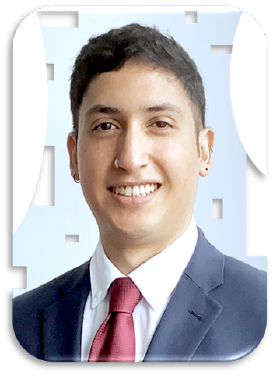

**Alan A. Díaz Montiel** holds a Postdoctoral Fellowship at the Neural Systems and Brain Signal Processing Lab (NSBSPL) which is part of the Krembil Research Institute - University Health Network (UHN) and the University of Toronto (Toronto, Canada). He obtained a PhD in Computer Science from the University of Dublin, Trinity College (Dublin, Ireland) in 2021. His research focuses on the analysis of electrophysiological signals, mainly intracranial EEG, aiming to identify cross-regional functional subnetworks in the human brain that can serve as biomarkers for epilepsy and other neurological disorders, and cognitive processes. His expertise is in the areas of Telecommunications, Simulation Systems, Signal Processing, Data Science and Applied AI.

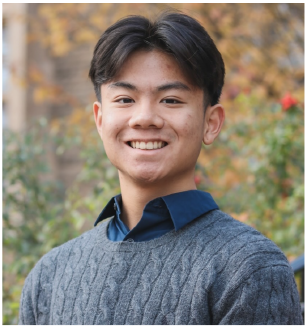

**Richard Zhang** is an Honours Bachelor of Health Sciences candidate at McMaster University and a research student at the Neural Systems and Brain Signal Processing Lab (NSBSPL). His work has involved analysis of high dimensional embeddings, graph neural network architectures, and unsupervised explainability techniques for machine-learning- transformed electrophysiological data. As an aspiring clinician scientist, Richard’s research interests include biomedical signal processing, machine learning explainability, and e-health bioinformatics.

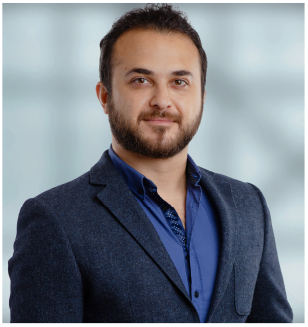

**Milad Lankarany** is a Scientist at the Krembil Research Institute - University Health Network, and an Assistant Professor at the Institute of Biomedical Engineering & Department of Physiology at the University of Toronto, Canada. Dr. Lankarany is the principal investigator at the Neural Systems and Brain Signal Processing Lab (NSBSPL). He obtained his PhD in Elec- trical and Computer Engneering from Concor- dia University, Montreal, Quebec, Canada. His research activities involve Computational Neu- rocience, Brain Signal Processing, Biologically-inspired AI, Deep Brain Stimulation, Neurotechnology and Wearable Systems.

## Notes

### Competing Interest Statement

The authors have declared no competing interest.

### Funding Statement

This study was partially supported by Dr. Lankarany's Krembil Seed Fund.

### Author Declarations

The study used ONLY openly available human data that were originally located at: 1) The OpenNeuro Epilepsy-iEEG-Multicenter-Dataset with accession number ds003029, available at https://openneuro.org/datasets/ds003029/versions/1.0.7; 2) The SWEC-ETHZ iEEG Database, available at http://ieeg-swez.ethz.ch/.

